# Knowledge and Awareness-based Survey of COVID-19 within the Eye Care Profession in Nepal: Misinformation is Hiding the Truth

**DOI:** 10.1101/2020.06.06.20123505

**Authors:** Sandip Sanyam Das, Sanjay Sah Kumar, Pankaj Chaudhary, Jeremy J Hoffman

## Abstract

**Background:** Nepal is currently under lockdown due to the COVID-19 pandemic with misinformation circulating on social media. This study aimed to analyse the knowledge and awareness of COVID-19 amongst eye care professionals in Nepal.

**Methodology:** We invited 600 participants from 12 ophthalmic centres across Nepal to complete a qualitative, anonymous online survey.

**Results:** Of the 600 eye care professionals invited, 310 (51%) participated in the survey. The symptoms of COVID-19 were known to 94%, whilst only 49% were aware of disease transmission. 98% of participants recognised the World Health Organization’s (WHO) awareness message, yet 41% of participants felt that consumption of hot drinks helps to destroy the virus. 41% disagreed that PPE should be mandatory for eye care practitioners.

**Conclusion:** There is still considerable scope to improve the knowledge of COVID-19 amongst ophthalmic professionals in Nepal. Opinion is also split on measures to prevent transmission, with misinformation potentially fuelling confusion.

## Introduction

The first case of COVID-19 in Nepal was reported on 13^th^ January 2020;^1^ currently the total count in Nepal has already passed 2000. In late 2019, an increase in cases of pneumonia of unknown aetiology in Wuhan, Hubei Province in China, prompted intense research.^2^ The Chinese Center for Disease Control and Prevention isolated and confirmed this pathogen as a novel type of coronavirus through a throat swab. This coronavirus was subsequently named 2019 novel-Coronavirus by the World Health Organization (WHO),^3^ who suggested on 25^th^ Dec 2019 that the virus is less contagious to healthcare professionals compared to other recent coronaviruses responsible for severe acute respiratory syndrome (SARS) and the Middle East respiratory syndrome (MERS).^4^ The virus was subsequently renamed as SARS-CoV-2 by the International Committee of Taxonomy of Viruses (ICTV),^5^ with the disease it causes named coronavirus disease 2019 (COVID-19). On 31 January 2020, the WHO characterised the outbreak as a public health emergency of international concern.^6^

Although the exact origin of the virus still remains elusive, the natural host of the virus is believed to be a species of bat.^7^ Initially, it was thought to be transmitted only as a zoonotic infection from animal to human, but subsequent research demonstrated that SARS-CoV-2 is highly contagious and can rapidly transmit between people through respiratory droplets (> 5-10 µm in diameter) and fomites.^2, 8–12,13^ The virus can remain viable on different surfaces, which may become a source of infection to individuals if hands are not washed properly.^14, 15^

COVID-19 varies in severity from being asymptomatic to causing respiratory failure that necessitates mechanical ventilation and support in an intensive care unit (ICU). Severe disease can lead to multiorgan failure, coma, and death.^16^ COVID-19 has been previously reported to be associated with conjunctivitis in humans.^17^

Developing countries such as Nepal, with limited healthcare infrastructure and capacity, may struggle to cope if the pandemic follows a similar trajectory to other countries. The Government of Nepal implemented a nationwide “lockdown” since the declaration of the pandemic on 23^rd^ March 2020, with severe limitations on movement of people. During this period, there have been many rumours and misinformation on COVID-19 emerging. The aim of this study was to review the knowledge and awareness of COVID-19 amongst eye care personnel in Nepal.

## Methodology

We performed a qualitative approach to quantify knowledge and awareness of COVID-19 amongst eye care personnel working in Nepal. This study conformed to the tenets of the Declaration of Helsinki, with a mandatory consent form that had to be completed before commencing the survey. As no patients were involved in this study, ethical permission was not required from our organisation. The research was carried out as an online questionnaire completed between 28^th^ March to 18^th^ April 2020. Nepal has approximately 1600 eye care professionals of different cadres (ophthalmologists, optometrists, ophthalmic assistants, and other ophthalmic personnel). The survey was sent to 600 of these clinicians working in Nepal. Participation was anonymous and voluntary. Representatives from a random selection of 12 of the 18 secondary or tertiary eye hospitals in Nepal were contacted and asked to select 50 individuals at their institution randomly, who were then invited to complete the survey.

The survey was divided into two sections: section 1 – demographic information; section 2 - 25 open- and closed-ended questions relating to knowledge and awareness of COVID-19 developed by a focused group of statisticians, microbiologists, research optometrists, and ophthalmologists. We implemented a non-probability convenience sampling method to reach the study population through social media applications (WhatsApp, Facebook, Telegram, Email and Viber). Each participant had only one chance to take the survey and two reminders were sent for individuals to complete the survey. The responses were anonymous, with no participant-identifiable questions. Data were exported into MS Excel. An overall performance score was calculated from the average of the 12 Knowledge questions for all the participants. These were categorised as “excellent” (if the average of correct responses was 80% or above), “satisfactory” (if the average of correct responses was 65% or above), or “poor” (if the average of correct responses was below 55%), in a similar way to academic grading.

## Results

The survey was completed by 310 (51%) individuals (55.5% male). The majority of participants (83.9%) were in age group 20-30. The demographic details are presented in Table 1.

**Table 1:**
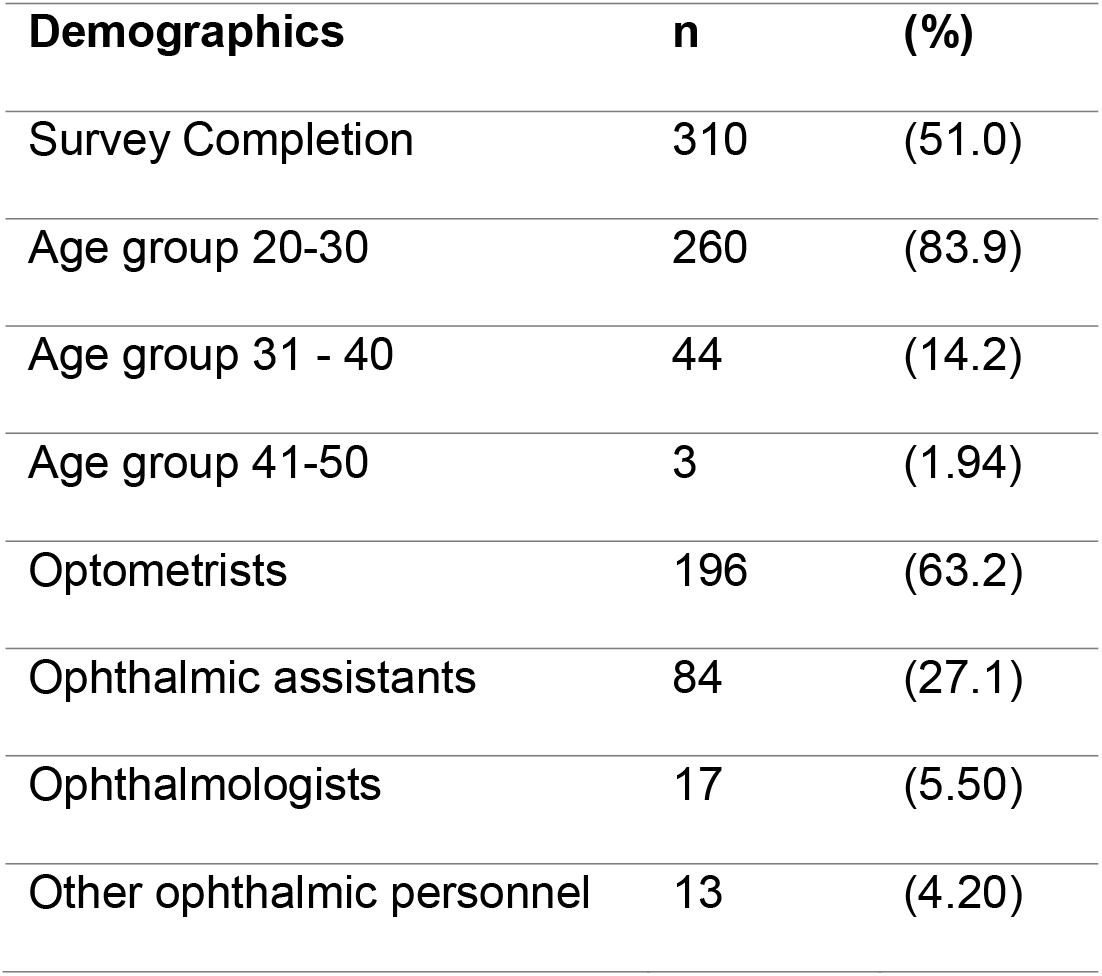
Demographic Details of Participants.

The responses to questions are given in Table 2.

**Table 2.**
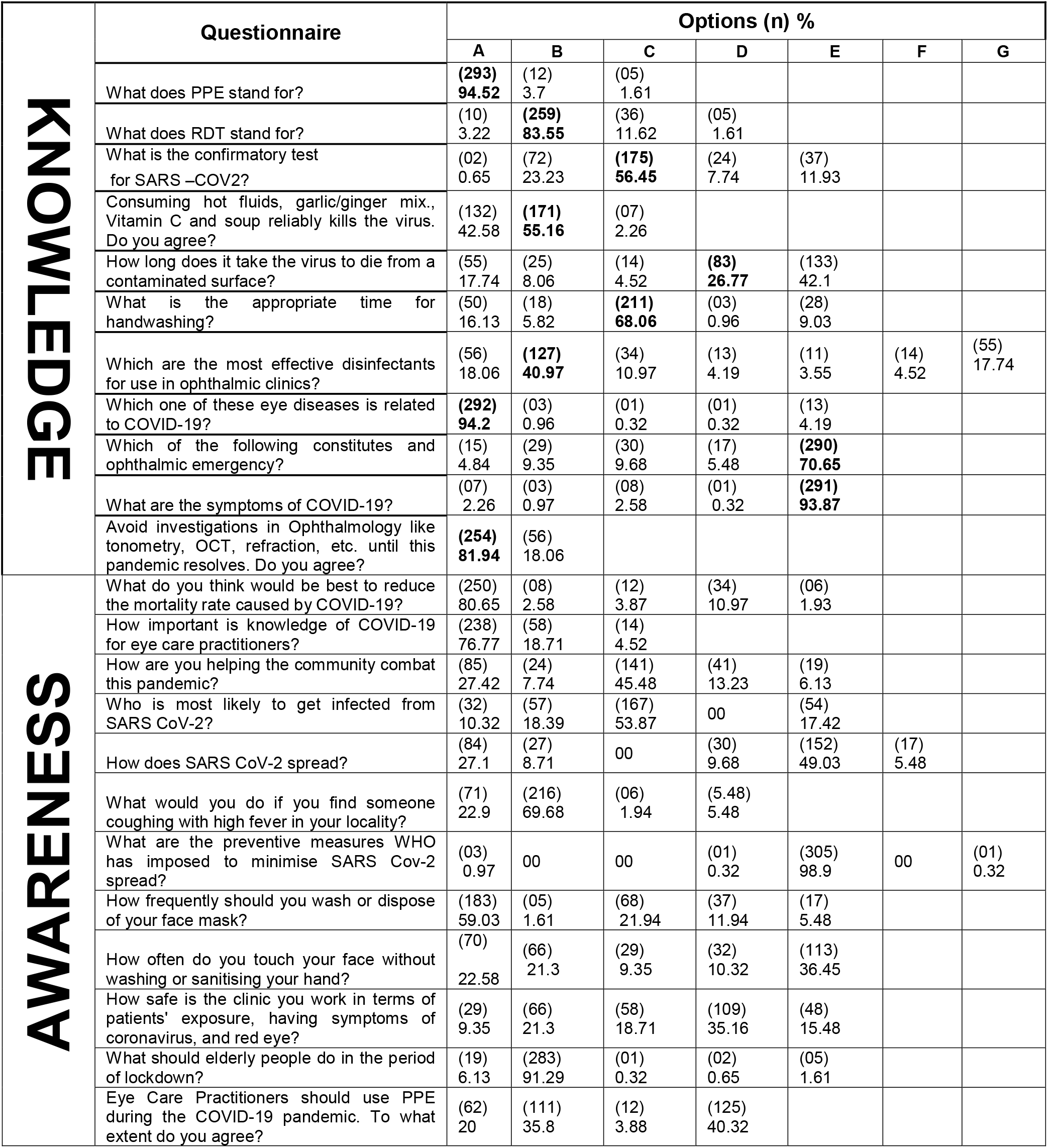
Knowledge and Awareness Questionnaire. Correct answers are given in bold. The number of possible responses differed between questions. The number of responses are given in brackets, with percentages below. The order of questions given here, separating questions assessing knowledge and awareness, differs to those presented to participants. Please see Supplemental Material for the full questions and as presented to participants with answers as

Of the participants, 76% said that knowledge of COVID-19 is essential to eye care practitioners. Despite this, there was a significant variation in the percentage of questions answered correctly. The symptoms of COVID-19 were known to 94%. However, only 54% were aware that anyone can be infected with SARS-CoV-2 and only 49% of the participants were aware of how COVID-19 is spread. Only 5% of the participants did not know the abbreviation of PPE (personal protective equipment) and 17% were unaware of the abbreviation RDT (rapid diagnostic tests). Regarding how COVID-19 is diagnosed, 56% responded correctly that Reverse Transcriptase-PCR for SARS-CoV-2 is used. Almost 98% of participants recognised the WHO’s awareness message, although 41% of participants believed that consumption of hot drinks kills the virus (in contradiction to information from the WHO). 80% of the participants felt that social distancing would be the key to restrict the disease spread.

Almost half of the participants were not sure regarding the life span of the virus on different surfaces, whilst 41% disagreed on a point that PPE is should be mandatory for eye care practitioners. 20% of the participants were not aware on the correct maintenance of a mask, which is an essential component of PPE. 59% of participants had PPE to work under COVID-19 crisis. The majority of participants were aware that washing hands with soap and water or alcohol-based sanitiser for at least 20 seconds kills viruses. Only 41% of the participants were able to answer that the multipurpose disinfectant used in clinics was 70% ethanol. 45% of the participants said that they follow WHO COVID-19 guidelines.

Interestingly, 94% of the participants knew that the most reported ocular manifestation of SARS-CoV-2 is conjunctivitis. Despite being ophthalmic clinicians, 30% were unable to identify what constituted an ocular emergency. 82% of the participants believed that diagnostic testing like tonometry, ocular coherence tomography, etc. must be postponed.

Altogether 81.9% (n = 254) of participants agreed with the statement ‘Avoid investigations in ophthalmology such as tonometry, OCT, refraction, etc. until this pandemic resolves. Descriptive reasons for allowing routine ophthalmic testing to continue were grouped into three categories: 1) if necessary in case of emergency 19/56 (33.93%); 2) perform only with COVID-19 infection control measures to aid in diagnosis 26/56 (46.43%); and 3) should perform as usual for routine attendance regardless of the COVID-19 situation 11/56 (19.64%).

The free text question ‘Being an eye care practitioner, what do you think is wise to do during this period of lockdown?’ The descriptive answers were categorised into different sub-categories, illustrated in Figure 1.

**Figure 1:**
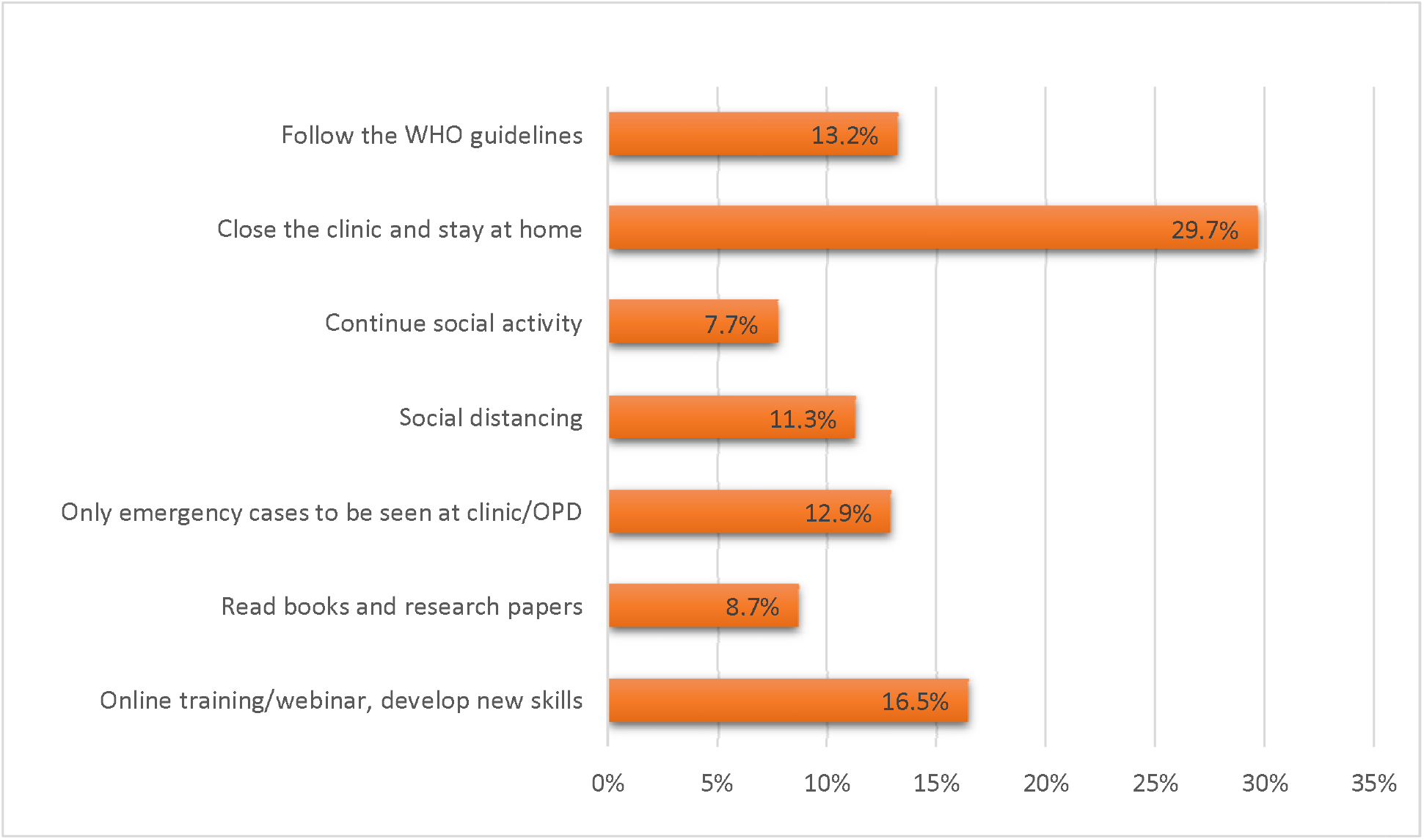
What should ophthalmic professionals do during lockdown in Nepal?

The calculated knowledge performance score was 69.75%, falling into the “satisfactory” category.

## Discussion

In Nepal, COVID-19 has been in the headlines since the beginning of 2020, and at the forefront of people’s minds since the lockdown began in March 2020. Currently, there are sporadic cases within the country according to the WHO.^18^ The Government of Nepal, in partnership with the WHO, is working tirelessly in preventing and controlling the disease. Building awareness around hygiene measures (hand wash procedure, respiratory hygiene, etc.) and social distancing have been given priority. Despite these evidence-based messages, there has been a considerable amount of misinformation and myths circulating within the country.

Our survey shows that there is considerable ignorance amongst those questioned regarding COVID-19. Only 56% felt it mandatory to use PPE if one is working in the ophthalmology/optometry clinic, despite reports of fatalities of ophthalmic clinicians due to COVID-19.^19^ The Centre for Disease Control and Prevention (CDC) and WHO have created guidelines for the safe running of emergency care in an ophthalmic setting. They have emphasised that PPE must be mandatory for ophthalmic clinicians to reduce the risk of contracting COVID-19.^20^ The PPE in the guidelines includes essentials to cover the mouth, nose (N95 mask preferably), eyes (goggles), face (visors), hands (gloves), and breath shields attached to the slit lamp.^21^ A similar proportion of participants surveyed (59%) had their clinics equipped with these safety measures. This suggests there are still challenges with regards to the provision of adequate PPE, a similar picture to that seen globally.

Half of the participants were not aware that the detection of novel SARS Cov-2 was confirmed by RT-PCR.^22^ One third of the participants felt that drinking things such as hot water, ginger-garlic mix, vitamin C or soup kills the virus directly or indirectly. There is no evidence to support this statement, which has been frequently circulating around social media as misinformation.^23, 24^

Approximately half of the participants were unaware of the lifespan of the virus on different surfaces.^13, 14^ Knowledge of this can help clinicians to appropriately disinfect surfaces and equipment. Several participants were not sure if they should clean their masks daily, while others did not wear masks, and a few washed it once every two days. The WHO recommends using a mask whilst in clinic, and importantly, there is a high risk of the infection being transmitted if the mask is not disposed of or sterilised properly on a daily basis, especially in susceptible areas.^15, 25^

The CDC and WHO recommends at least 20 seconds should be spent while washing hands.^26^. Most participants said they follow this recommendation.

The CDC recommends 70% ethanol to be used as a multipurpose disinfectant in ophthalmology clinics and other surface cleaning activities.^22^ More than half of participants (59.0% (n = 183)) believed other less-potent disinfectants were adequate, which highlights an area that can be improved on by further education of clinic staff.

Cases of conjunctivitis have been reported.^17^ Conjunctival secretions and tears from infected patients may contain viral RNA, whilst those with conjunctival symptoms may pose a higher risk in terms of transmission.^27^ However, it is a rare symptom of COVID-19. In a recent study, of the total reported cases of ocular manifestations of COVID-19, only 0.8% had conjunctivitis; this is in keeping with other coronaviruses which are also known to (infrequently) cause conjunctivitis.^12, 28^ Clinicians therefore need to be cautious while examining conjunctivitis patients as there is a risk they may be sources of SARS-CoV-2 infection. Knowledge of this is therefore important to ophthalmic practitioners. The majority of the participants knew that conjunctivitis was the most common ocular manifestation of COVID-19, which is reassuring.

As a knowledge assessment on ocular emergency within the context of COVID-19, 5% of the participants felt blurred vision was an emergency, which in our view unless acute in onset, would not be classed as such. There is therefore a chance that routine patients are being seen unnecessarily during lockdown by a minority of clinicians.

A free-text opinion was taken from the participants on what should be done by a professional during lockdown, to gauge personal behaviour and frustrations they may feel. The responses to this question are given in Figure 1. Interestingly, the majority felt that they should not be working and should self-isolate at home. At the other end of the spectrum, 7.7% still felt that social activities should continue. This suggests their opinions are somewhat divided.

Approximately half of participants believed that only immunocompromised individuals and/or elderly people are susceptible to the SARS-CoV-2, whereas half recognised that anybody can be infected, as stated by WHO.^29^ There were mixed opinions regarding the spread of SARS-CoV-2, with approximately 10% of participants feeling that SARS-CoV-2 is transmitted as an aerosol. This mirrors the literature, as it still remains unclear exactly how the virus spreads within the environment. A recent study of aerosols did not find any contamination of air with SARS-CoV-2 in an isolation ward of COVID-19 patients.^13^

There are some potential limitations to this study. Although we selected participants at random from the main eye hospitals in Nepal, only just over half of all those invited responded to the survey. This may cause a degree of selection bias. Furthermore, the COVID-19 pandemic is a rapidly evolving situation, which means that some of the “correct” answers to our survey that were accurate at the time of questioning, may have changed by the time of analysis. The strength of this study is that this is the only such study to have been conducted in Nepal, within a short timeframe.

## Conclusion

Our study shows that knowledge amongst eye care practitioners is classed as satisfactory. We therefore recommend ophthalmic practitioners to enhance and develop this knowledge as a priority. There are abundant, free webinars on COVID-19 and eye care from which individuals can benefit. The views expressed within the awareness questions are concerning, as some of these responses highlight a degree of misinformation, which may be of concern for institutions and local government. Further work must be carried out by employers, as well as local and national governments in Nepal, in order to prevent the spread of misinformation, and support practitioners to carry out eye care during this pandemic safely. The views expressed in this study reflect those of educated professionals: it is therefore likely that public awareness is less, with belief in rumours greater, than what we have identified. Further studies are necessary to accurately assess the opinions of the general public on COVID-19 in Nepal.

With rumours spreading quickly during crises, there has been an increasing amount of “fake news” or misinformation circulating in Nepal during this pandemic, and this study highlights some of these issues. It is crucial that clinicians remember to check the scientific evidence before altering their practice or recommending interventions to their patients.

### Contributing authorship statement

SD, SK, and PC conceived the design. SD and SK collected the data. All authors analysed the data and have approved the final manuscript.

### Conflict of Interest

Authors do not have any conflict of interest to declare.

### Funding

No funding was received for this study.

### Data sharing statement

Data can be shared on reasonable request to the corresponding author by bone fide researchers.

## Data Availability

Data can be made available on relevant request to the corresponding author.

## Notes

### Competing Interest Statement

The authors have declared no competing interest.

### Funding Statement

No funding received

### Author Declarations

Sagarmatha Chaudhary Eye Hospital Ethical Board

